# Visible and real sizes of the COVID-19 pandemic in Ukraine

**DOI:** 10.1101/2021.03.19.21253938

**Authors:** Igor Nesteruk

**Affiliations:** Institute of Hydromechanics, National Academy of Sciences of Ukraine, Zheliabova 8/4, UA-03680 Kyiv, Ukraine; National Technical University of Ukraine “Igor Sikorsky Kyiv Polytechnic Institute”, Prosp.Peremohy 37, UA-03056, Kyiv, Ukraine

**Keywords:** COVID-19 pandemic, epidemic dynamics in Ukraine, mathematical modeling of infection diseases, SIR model, parameter identification, statistical methods

## Abstract

To simulate how the number of COVID-19 cases increases versus time, various data sets and different mathematical models can be used. In particular, previous simulations of the COVID-19 epidemic dynamics in Ukraine were based on smoothing of the dependence of the number of cases on time and the generalized SIR (susceptible-infected-removed) model. Since real number of cases is much higher than the official numbers of laboratory confirmed ones, there is a need to assess the degree of data incompleteness and correct the relevant forecasts. We have improved the method of estimating the unknown parameters of the generalized SIR model and calculated the optimal values of the parameters. It turned out that the real number of diseases exceeded the officially registered values by about 4.1 times at the end of 2020 in Ukraine. This fact requires a reassessment of the COVID-19 pandemic dynamics in other countries and clarification of world forecasts.

## Introduction

The studies of the COVID-19 pandemic dynamics are complicated by incomplete information about the number of patients (e.g., reported by WHO [1]), a very large percentage of whom are asymptomatic. In the early stages of the pandemic, there was also a lack of tests and knowledge about the specifics of the infection spread. Because of this, there are more and more evidences of COVID-19 patient appearances before the first officially-confirmed cases [2-6]. These hidden periods of the epidemics in different countries and regions were estimated in [7-11] with use of the classical SIR model [12-14] and the statistics-based method of the parameter identification developed in [15, 16]. In particular, first COVID-19 cases probably have appeared already in August 2019 [9-11].

For Ukraine, different simulation and comparison methods were based on official accumulated number of laboratory confirmed cases [17, 18] (these figures coincided with the official WHO data sets [1], but WHO stopped to provide the daily information in August 2020) and the data reported by Johns Hopkins University (JHU) [19]. In particular simple comparisons of epidemic outbreaks in Ukraine neighboring countries can be found in [20-22]. The classical SIR model was used in [7-11, 23-25]. The weakening of quarantine restrictions, changes in the social behavior and the coronavirus activity caused changes in the epidemic dynamics and corresponding parameters of models. To detect and simulate these new epidemic waves, a simple method of numerical differentiations of the smoothed number of cases and generalized SIR model were proposed and used in [11, 26-30]. In particular, nine epidemic waves in Ukraine were calculated [11, 26-30]. Since the Ukrainian national statistics does not look complete (see, e.g., the results of total staff testing in two schools and two children gardens in the Ukrainian city of Chelnytskii, [31]), there is a need to assess the extent of this incompleteness and determine the true size of the COVID-19 epidemic in Ukraine, which became the subject of this article.

### Data

We will use the data set regarding the accumulated numbers of confirmed COVID-19 cases in Ukraine from national sources [17, 18]. The corresponding numbers *V*_*j*_ and moments of time *t*_*j*_ (measured in days) are shown in the supplementary Table A. It must be noted that this table does not show all the COVID-19 cases occurred in Ukraine. Many infected persons are not identified, since they have no symptoms. For example, employees of two kindergartens and two schools in the Ukrainian city of Chmelnytskii were tested for antibodies to COVID-19, [31]. In total 292 people work in the surveyed institutions. Some of the staff had already fallen ill with COVID-19 or were hospitalized. Therefore, they were tested and registered accordingly. In the remaining tested 241 educators, antibodies were detected in 148. Therefore, the number of identified patients (51) in these randomly selected institutions was 3.9 times less than the actual number (51+148) of COVID-19 cases. Many people know that they are ill, since they have similar symptoms as other members of families, but avoid making tests. Unfortunately, one laboratory confirmed case can correspond to several other cases which are not confirmed and displayed in the official statistics. The number of cases in Ukraine reported by COVID-19 Data Repository by the Center for Systems Science and Engineering (CSSE) at Johns Hopkins University (JHU) [19] is 2-3 % higher than the Ukrainian national statistics [17, 18] yields (see [30]). Nevertheless, the special simulations will demonstrate a significant incompleteness of both data sets.

### Generalized SIR model

The classical SIR model for an infectious disease [12-14] was generalized in [11, 27-30] to simulate different epidemic waves. We suppose that the SIR model parameters are constant for every epidemic wave, i.e. for the time periods: 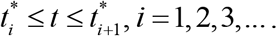 Than for every wave we can use the equations, similar to [12-14]:

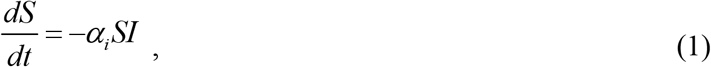

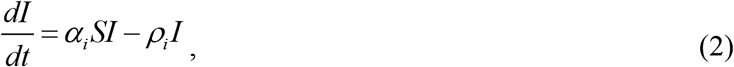

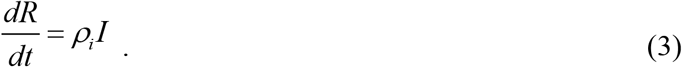

Here *S* is the number of susceptible persons (who are sensitive to the pathogen and **not protected**); *I* is the number of infected persons (who are sick and **spread the infection)**; and *R* is the number of removed persons (who **no longer spread the infection**; this number is the sum of isolated, recovered, dead, and infected people who left the region). Parameters *α*_*i*_ and *ρ*_*i*_ are supposed to be constant for every epidemic wave.

To determine the initial conditions for the set of equations (1)–(3), let us suppose that at the beginning of every epidemic wave 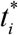:

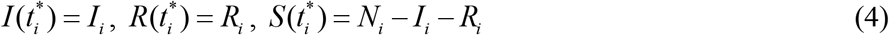

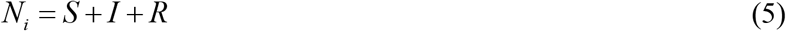

In [11, 27-30] the set of differential equations (1)-(3) was solved by introducing the function

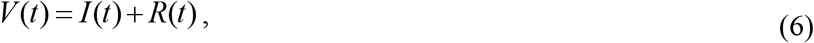

corresponding to the number of victims or the cumulative confirmed number of cases. For many epidemics (including the COVID-19 pandemic) we cannot observe dependencies *S*(*t*), *I*(*t*) and *R*(*t*) but observations of the accumulated number of cases *V*_*j*_ corresponding to the moments of time *t*_*j*_ provide information for direct assessments of the dependence *V* (*t*). The corresponding analytical formulas for this exact solution can be written as follows:

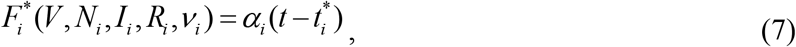

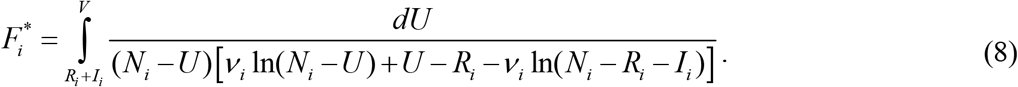

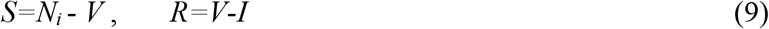

Thus, for every set of parameters *N*_*i*_, *I*_*i*_, *R*_*i*_, *ν*_*i*_ and a fixed value of *V*, integral (8) can be calculated and the corresponding moment of time can be determined from (7). Then functions *I(t)* and *R(t)* can be easily calculated with the use of formulas (9). The saturation levels *S*_*i∞*_; *V*_*i∞*_= *N*_*i*_− *S*_*i∞*_ (corresponding the infinite time moment) and the final day of the *i-th* epidemic wave (corresponding the moment of time when the number of persons spreading the infection will be less then 1) can be calculated with the use of equations available in [11, 27-30].

### Parameter identification procedure

In the case of a new epidemic, the values of its parameters are unknown and must be identified with the use of limited data sets. For the second and next epidemic waves (*i* > 1), the moments of time 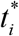 corresponding to their beginning are known. Therefore the exact solution (7)-(9) depend only on five parameters - *N*_*i*_, *I*_*i*_, *R*_*i*_,*ν*_*i*_, *α*_*i*_, when the registered number of victims *V*_*j*_ is the random realization of its theoretical dependence (6). If we assume, that data set *V*_*j*_ is incomplete and there is a constant coefficient *β*_*i*_ ≥ 1, relating the registered and real number of cases during the *i-th* epidemic wave:

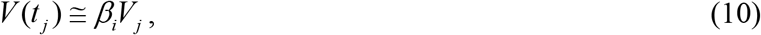

the number of unknown parameters increases by one.

Then the values *V*_*j*_, corresponding to the moments of time *t*_*j*_ and relationship (10) can be used in eq. (8) in order to calculate 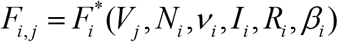 for every fixed values of *β*_*i*_, *N*_*i*_, *ν*_*i*_, *I*_*i*_, *R*_*i*_ and then to check how the registered points fit the linear dependence (7) which can be rewritten as follows:

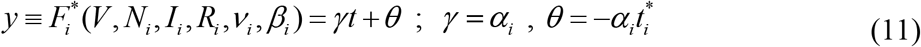

We can calculate the parameters *γ* and *θ*, by treating the values 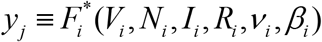 and corresponding time moments *t*_*j*_ as random variables. Then we can use the observations of the accumulated number of cases and the linear regression in order to calculate the coefficients 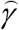 and 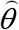 of the regression line

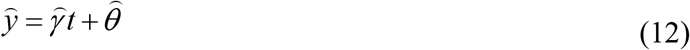

using the standard formulas (see, e.g., [32]). Values 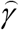 and 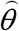 can be treated as statistics-based estimations for parameters *γ* and *θ* from relationships (11).

The reliability of the method can be checked by calculating the correlation coefficients *r*_*i*_ (see e.g., [13]) for every epidemic wave and checking how close are their values is to unity. We can use also the F-test [13] for the null hypothesis that says that the proposed linear relationship (11) fits the data set. Similar approach was used in [7-11, 15, 16, 23-30, 33, 34]. To calculate the optimal values of parameters *β*_*i*_, *N*_*i*_, *ν*_*i*_, *I*_*i*_, *R*_*i*_, we have to find the maximum of the correlation coefficient for the linear dependence (11).

The exact solution (7)-(9) allows avoiding numerical solutions of differential equations (1)-(3) and significantly reduces the time spent on calculations. A new algorithm [11, 29, 30] allows estimating the optimal values of SIR parameters for the *i-th* epidemic wave directly (without simulations of the previous waves). To reduce the number of unknown parameters, we can use the relationship *V*_*i*_= *I*_*i*_+ *R*_*i*_ which follows from (6) and (10). To estimate the value *V*_*i*_, we can use the smoothed accumulated number of cases [11, 26-30]

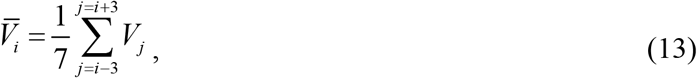

and the relationship 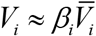 following from (10) (*i* corresponds to the moment of time 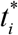). One more relationship

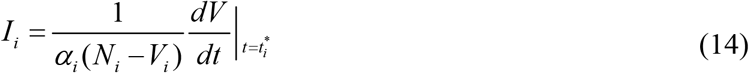

can be obtained with the use of (5) and formula

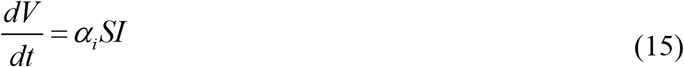

(following from (2) and (3)). To estimate the average number of new cases *dV/dt* at the moment of time 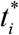 in eq. (14), we can use the numerical differentiation of (13):

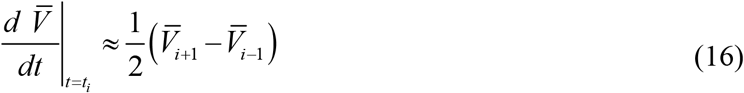

and relationship (10). Thus we have only three independent parameters *β*_*i*_, *N*_*i*_and *ν*_*i*_. To calculate the value of parameter *α*_*i*_, some iterations can be used (see details in [11]).

## Results

The optimal values of parameters and other characteristics of the ninth COVID-19 pandemic wave in Ukraine are listed in Table 1 for two cases. For SIR simulations we have used the same period of time *T*_*c*_: December 11-24, 2020 and corresponding values of *V*_*j*_ and *t*_*j*_ from Table A. In the first case we assumed that the numbers of registered cases coincide with the real one (*β*_9_ =1). A similar SIR simulation of the 9th epidemic wave in Ukraine has already been reported in [30], but now we have managed to find a new (larger in value) maximum of the correlation coefficient.

**Figure.**
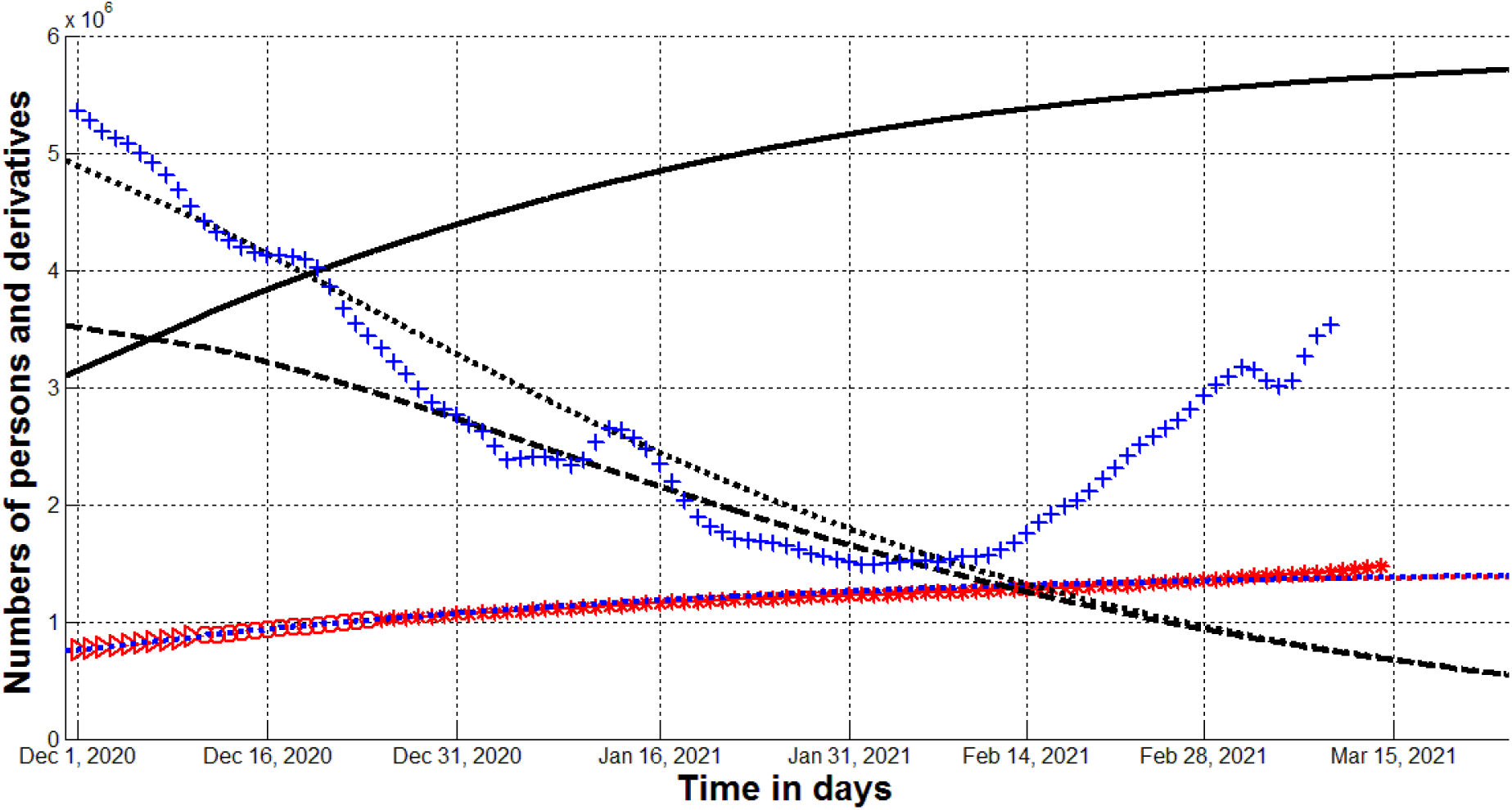
Visible (red) and real (black) COVID-19 epidemic dynamics in Ukraine. Red markers show accumulated numbers of cases *V*_*j*_ from Table A. “Circles” correspond to the accumulated numbers of cases taken for calculations (during period of time *T*_*c*_); “triangles” – numbers of cases before *T*_*c*_; “stars” – number of cases after *T*_*c*_. Blue and black colors correspond to the case *β*_9_ =4.1024; the blue “crosses” show derivative (16) multiplied by 100*β*_9_; the blue dotted line represents dependence *I* (*t*) / *β*_9_. Numbers of victims *V(t)=I(t)+R(t)* – black solid lines; numbers of infected and spreading *I(t)* multiplied by 5 – dashed; derivatives *dV/dt* (eq. (15)) multiplied by 100 – dotted. The red dotted line represents dependence *I* (*t*) / *β*_9_ for the case *β*_9_ =1.

**Table 1.** **Calculated optimal values of SIR parameters and other characteristics of the ninth COVID-19 pandemic wave in Ukraine.**

| Characteristics | 9<sup>th</sup> epidemic wave,<br>$i=9$,<br>$\beta_i=1$(without<br>optimization) | 9<sup>th</sup> epidemic wave,<br>$i=9$,<br>$\beta_i=4.1024$ |
| --- | --- | --- |
| $I_i$ | 148,390.742887927 | 668,766.512528977 |
| $R_i$ | 732,364.542826359 | 2,944,443.97158531 |
| $N_i$ | 2,960,000 | 12,307,200 |
| $\nu_i$ | 2,275,096.81932990 | 9626720.00517470 |
| $\alpha_i$ | 3.48830042313868e-08 | 7.59399763733452e-09 |
| $\rho_i$ | 0.0793622119754996 | 0.0731052889745776 |
| $1/\rho_i$ | 12.6004552432172 | 13.6789008569236 |
| $r_i$ | 0,998201733486790 | 0.998205208046402 |
| $S_{i\infty}$ | 1,530,454 | 6,372,870 |
| $V_{i\infty}$ | 1,429,546 | 5,934,330 |
| Final day of the<br>epidemic wave | April 17, 2022 | July 24, 2022 |

The last column of Table 1 illustrate the results of SIR simulations with the non-prescribed value

of *β*_9_. The maximum of the correlation coefficient was achieved at *β*_9_ =4.1024. This result testifies that the main part of the epidemic in Ukraine is invisible. At the end of 2020 the real numbers of COVID-19 cases probably were more than 4 times higher than registered ones. The real final size of the ninth epidemic wave *V*_9∞_is expected to be around 6 million. Unfortunately, we cannot wait for the end of the pandemic before the summer of 2022 (if vaccinations will not change this sad trend).

Knowing the optimal values of parameters, the corresponding SIR curves can be easily calculated with the use of exact solution (7)-(9) and compared with the pandemic observations after *T*_*c*_. The results are shown in Figure by different colors. Black and blue lines and markers correspond to the case *β*_9_ =4.1024. The solid black line shows complete accumulated number of cases (visible and invisible); the dashed line represents the complete number of infected persons multiplied by 5, i.e. *I(t)x5*; dotted black line represent the derivative *dV/dt* (which is an estimation of the real daily number of new cases) calculated with the use of (15) and multiplied by 100. The red dotted line shows the dependence *V(t)* for the case *β*_9_ =1 (assuming that all the cases are registered). The red “circles”, “triangles”, and “stars” correspond to the accumulated numbers of cases registered during period of time taken for SIR simulations *T*_*c*_, before *T*_*c*_, and after *T*_*c*_, respectively (taken from Table A). The blue dotted line represents dependence *I* (*t*) / *β*_9_. The blue crosses show the estimation of the derivative (16) multiplied by 100*β*_9_.

## Discussion

According to the results of our study, we can only say that in the case of suitability of the generalized SIR model, the value *β*_9_ =4.1024 and other optimal values of its parameters (given in the last column of Table 1) are the most reliable (provide the maximum value of the correlation coefficient). Therefore, we used additional methods to verify the calculations and showed some results in Fig. The blue dotted line represents dependence *I* (*t*) / *β*_9_ which must be close to the registered number of cases (red markers). The coincidence is very good. Significant deviations began to appear only in March 2021, which can be explained by the beginning of the next (tenth) epidemic wave in Ukraine. The blue crosses show the estimation of the real daily number of new cases (derivative (16) multiplied by 100*β*_9_) and have to be close to the black dotted line. Significant deviations began to appear only in mid-February 2021, which can be explained canceling the lockdown on January 24, 2021. May be the beginning of the tenth epidemic wave is connected also with the starting the lessons at schools and universities and mutations of the coronavirus.

The calculated coefficient of epidemic visibility *β*_9_ =4.1024 correlates with the results of testing employees of two kindergartens and two schools in the Ukrainian city of Chmelnytskii [31] which revealed the value 3.9. Probably that large discrepancy between registered and actual number of cases occurred not only in Ukraine. For example, total testing in Slovakia (65.5% of population was tested on October 31-November 1, 2020) revealed a number of previously undetected cases, equal to about 1% of the population [35]. On November 7 next 24% of the population was tested and found 0.63% of those infected [36]. According to the WHO report at the end of October, the number of detected cases in Slovakia was also approximately 1% of population [1].

Many authors are and will be trying to predict the COVID-19 pandemic dynamics in many countries and regions [7-11, 16, 23-30, 37-102]. The results of this study indicate that reliable estimates of its dynamics require consideration of incomplete data and constant changes of the conditions (quarantine restrictions, social distancing, coronavirus mutations, etc.).

## Data Availability

Data are in the Text

## Acknowledgements

The author is grateful to Oleksii Rodionov for his help in collecting and processing data.

## Supplementary materials

**Table A.** **Cumulative numbers of laboratory confirmed Covid-19 cases in Ukraine *V*_*j*_ according to the national statistics, [7, 8].**

| Day in<br>Decem-<br>ber<br>2020,<br>$t_j$ | Number<br>of cases,<br>$V_j$ | | Day in<br>Janu-<br>ary<br>2021 | Number of<br>cases,<br>$V_j$ | | Day in<br>Febru-<br>ary<br>2021 | Number of<br>cases,<br>$V_j$ | | Day in<br>March<br>2021 | Number of<br>cases,<br>$V_j$ |
| --- | --- | --- | --- | --- | --- | --- | --- | --- | --- | --- |
| 1 | 758264 |  | 1 | 1069517 |  | 1 | 1223879 |  | 1 | 1357470 |
| 2 | 772760 |  | 2 | 1074093 |  | 2 | 1227164 |  | 2 | 1364705 |
| 3 | 787891 |  | 3 | 1078251 |  | 3 | 1232246 |  | 3 | 1374762 |
| 4 | 801716 |  | 4 | 1083585 |  | 4 | 1237169 |  | 4 | 1384917 |
| 5 | 813306 |  | 5 | 1090496 |  | 5 | 1241479 |  | 5 | 1394061 |
| 6 | 821947 |  | 6 | 1099493 |  | 6 | 1244849 |  | 6 | 1401228 |
| 7 | 832758 |  | 7 | 1105169 |  | 7 | 1246990 |  | 7 | 1406800 |
| 8 | 845343 |  | 8 | 1110015 |  | 8 | 1249646 |  | 8 | 1410061 |
| 9 | 858714 |  | 9 | 1115026 |  | 9 | 1253055 |  | 9 | 1416438 |
| 10 | 872228 |  | 10 | 1119314 |  | 10 | 1258094 |  | 10 | 1425522 |
| 11 | 885039 |  | 11 | 1124430 |  | 11 | 1262867 |  | 11 | 1438468 |
| 12 | 894215 |  | 12 | 1130839 |  | 12 | 1268049 |  | 12 | 1451744 |
| 13 | 900666 |  | 13 | 1138764 |  | 13 | 1271143 |  | 13 | 1460756 |
| 14 | 909082 |  | 14 | 1146963 |  | 14 | 1273475 |  | 14 | 1467548 |
| 15 | 919704 |  | 15 | 1154692 |  | 15 | 1276618 |  | - | - |
| 16 | 931751 |  | 16 | 1160682 |  | 16 | 1280904 |  | - | - |
| 17 | 944381 |  | 17 | 1163716 |  | 17 | 1287141 |  | - | - |
| 18 | 956123 |  | 18 | 1167655 |  | 18 | 1293672 |  | - | - |
| 19 | 964448 |  | 19 | 1172038 |  | 19 | 1299967 |  | - | - |
| 20 | 970993 |  | 20 | 1177621 |  | 20 | 1304456 |  | - | - |
| 21 | 979506 |  | 21 | 1182969 |  | 21 | 1307662 |  | - | - |
| 22 | 989642 |  | 22 | 1187897 |  | 22 | 1311844 |  | - | - |
| 23 | 1001132 |  | 23 | 1191812 |  | 23 | 1317694 |  | - | - |
| 24 | 1012167 |  | 24 | 1194328 |  | 24 | 1325841 |  | - | - |
| 25 | 1019876 |  | 25 | 1197107 |  | 25 | 1333844 |  | - | - |
| 26 | 1025989 |  | 26 | 1200883 |  | 26 | 1342016 |  | - | - |
| 27 | 1030374 |  | 27 | 1206412 |  | 27 | 1347849 |  | - | - |
| 28 | 1037362 |  | 28 | 1211593 |  | 28 | 1352134 |  | - | - |
| 29 | 1045348 |  | 29 | 1216278 |  | - | - |  | - | - |
| 30 | 1055047 |  | 30 | 1219455 |  | - | - |  | - | - |
| 31 | 1064479 |  | 31 | 1221485 |  | - | - |  | - | - |

